# Excess mortality in India from June 2020 to June 2021 during the COVID pandemic: death registration, health facility deaths, and survey data

**DOI:** 10.1101/2021.07.20.21260872

**Authors:** Yashwant Deshmukh, Wilson Suraweera, Chinmay Tumbe, Aditi Bhowmick, Sankalp Sharma, Paul Novosad, Sze Hang Fu, Leslie Newcombe, Hellen Gelband, Patrick Brown, Prabhat Jha

**Author notes:** Address reprint requests to Professor Jha at CGHR, 30 Bond Street, Toronto, ON Canada M5B 2C8 or.

## Abstract

**Background:** India’s official death totals from the COVID pandemic are widely regarded as under-reports.

**Methods:** We quantified all-cause excess mortality in India, comparing deaths during the peak of the first and second COVID waves (Jul-Dec 2020 and April-June 2021) with month wise deaths in 2015-19 from three sources: Civil Registration System (CRS) mortality reports from 15 states or cities with 37% of India’s population; deaths in 0.2 million health facilities; and a representative survey of 0.14 million adults about COVID deaths.

**Results:** During the first viral wave, the median excess mortality compared to CRS baseline was 22% and 41%, respectively, in included states and cities, rising to 46% and 85% during the second wave. In settings with 10 or more months of data across the two waves, the median excess mortality was 32% and 37% for states and cities, respectively. Deaths in health facilities showed a 27% excess mortality from July 2020-May 2021, reaching 120% during April-May 2021. The national survey found 3.5% of adults reported a COVID death in their household in April-June 2021, approximately doubling the 3.2% expected overall deaths. The national survey showed 29-32% excess deaths from June 1, 2020 to June 27, 2021, most of which were likely to be COVID. This translates to 3.1-3.4 million COVID deaths (including 2.5-2.8 million during April-June 2021). National extrapolations from health facility and CRS data suggest 2.7-3.3 million deaths during the year.

**Conclusions:** India’s COVID death rate may be about 7-8 times higher than the officially reported 290/million population.

---

India has reported over 30 million cases of SARS-CoV-2, second only to the United States (US).^1^ India’s official cumulative COVID death count of 0.4 million implies a COVID death rate of approximately 290/million population, far lower than the UK or the US, each about 1900/million.^2^ India’s reported COVID death totals are widely believed to be under-reports,^3^ due to incomplete certification of COVID deaths, misattribution to chronic diseases, and because over 70% of India’s 10 million annual deaths occur in rural areas, often without medical attention.^4,5^ Thus, deaths are often not registered or medically certified.

Model-based estimates of cumulative COVID deaths in India range from a few hundred thousand to over four million,^6,7^ but models rely on official reports and apply varied assumptions. In the absence of near universal and timely death registration and release of data from India’s Sample Registration System (SRS), which tracks deaths in a random sample of about 1% of Indian homes,^8^ alternative approaches are needed to estimate COVID deaths. Excess all-cause mortality counts are recognized now by the World Health Organization (WHO) as an imperfect but useful such method.^9^

Here, we report on excess mortality in India using three data sources: 1) civil registration data from eight states and seven cities that comprise over a third of India’s population, 2) national facility-based death reporting, and 3) COVID mortality reported in a nationally representative telephone survey of 0.14 million adults.

## Methods

### Civil Registration System (CRS) data

India’s 1969 Registration of Births and Deaths Act requires compulsory registration of the facts of births and deaths (but not causes of death). A local registrar for each village in rural areas and for each municipality in urban areas is responsible for reporting, which is through electronic entry. The latest Civil Registration System (CRS) annual report is for 2019,^5^ but we obtained disaggregated data for 2020 and 2021 for this analysis from journalists and local NGOs, who received data through India’s Right to Information Act or through administrative requests. Our sample includes all states and cities where the proportion of registered deaths before 2020 covered at least 50% of expected deaths, yielding eight states (in decreasing population size: Madhya Pradesh, Tamil Nadu, Gujarat, Andhra Pradesh, Odisha, Kerala, Assam and Haryana) and seven cities (Ahmedabad, Gujarat; Mumbai and Nagpur, Maharashtra; Chennai, Tamil Nadu; Bengaluru, Karnataka; Hyderabad, Telangana; Kolkata, West Bengal). Table S1 in the Supplementary Appendix provides data for all states, including those not meeting the inclusion criteria. We report only aggregate deaths, as sex and socioeconomic-stratified data were incomplete.

### Facility-based death reporting

The Ministry of Health and Family Welfare tracks key outcomes relevant to national programs through an online Health Management Information System that covers about 0.2 million public hospitals and smaller facilities nationally, more than 90% of them rural.^10^ Data are updated monthly, including details on age group and broad cause of death (including “unknown cause”). During 2018-19, an average of 2.50 million annual deaths were reported, corresponding to approximately 52% of all facility-based deaths in the country.^8^

### Nationally representative survey

Cvoter India OmniBus is a nationally representative, computer-assisted telephone interview survey carried out daily for political and other socioeconomic purposes.^11^ It was adapted to report on COVID symptoms in March 2020, among adults aged 18 years or older, covering about 2100 randomly selected respondents weekly, drawn from nearly 4000 political ridings. Random digit dialing stratified for local mobile networks was used to sample India’s population, which has 90% or higher coverage with mobile phones.^12^ Cvoter participants are largely representative of the Indian population when compared to the 2011 census (Table S2 in Supplementary Appendix). The overall margin of error is +/- 3% at the national level and +/- 5% at the state level. The final data are weighted by age, gender, social group, education, voting or not in the last national election, and rural/urban locality.

The survey asks, “Have you seen flu-like symptoms like high fever, cold, dry cough, or similar symptoms in any family member (within your own household) or in your neighbourhood (people who you normally meet in your day-to-day life)ã” From June 2020, questions were added about COVID infection, hospitalization, or death, and vaccination and testing history. The mortality question asked, “Has anyone in your family or surroundings been infected from Corona Virusã” If the answer was yes, respondents were asked whether the infected individual died. From May 15, 2021, the age of death was ascertained. Teams of 125 callers worked from home to conduct weekly calls in 11 languages.

### Statistical methods

We examined CRS registered deaths in two periods: during the peak of the first viral wave (July-Dec 2020 in most places, varying somewhat by state and city)^13^ or the second viral wave (April-June 2021). We compared deaths in each wave to CRS deaths during previous years (2015-19 or the largest available subset of these years) for the same months to take into account seasonality. Because increases or decreases in registration, as well as delays in the registration process, may affect reported totals, we also compared registered deaths to the 2019 United Nations (UN) Population Division national death estimates, partitioned to states by rural/urban status based on the SRS proportions of 2016–18 average deaths (Table S3 in Supplementary Appendix).^4,14^ The UN demographic estimates adjust for the approximate 15% undercount in deaths in the SRS.^4,15^ Kerala, Gujarat, Bengaluru, and Hyderabad had slightly fewer expected than reported deaths compared to the baseline years of 2015-19. We assigned these as zero excess deaths, as it is improbable that total mortality fell during peak COVID weeks. For the subset of localities with 10 or more months of observations covering both viral waves, we summed the excess deaths and compared these to the UN demographic estimates. We applied similar procedures for the facility-based death reporting,^10^ comparing the average monthly reports for all-cause mortality from July-Dec 2020 and April-May 2021 to the average for these months for 2018-19.

We made two adjustments to the Cvoter results. First, we excluded deaths before age 35, which are unlikely to be from COVID^2^ (16.6% of reported deaths). Second, COVID deaths may have been over-reported because they were not restricted to immediate household members. To correct for this, we subtracted a background rate of 0.58% (observed from mid-February 2021 to April 2021, and during early June 2020, each period when confirmed COVID deaths were reported as below 200 daily nationally),^2^ assuming that 0.58% represents the background rate of adults who know of a COVID death among their wider social circles, but not in their household. We subject this assumption in a sensitivity analyses using 50% or 150% of the 0.58% used. We compared the adjusted COVID death proportions to expected deaths in a household, based on the 2019 UN total deaths (2020 or 2021 projected deaths yielded nearly identical results).^14^ Daily variation in expected deaths relied on Million Death Study data from 2004-14.^16^ Finally, we applied a cross-correlation statistic^17^ to compare the adjusted COVID deaths to confirmed COVID deaths, reported on covid19India.org,^2^ which compiles official and media reports daily. Analyses were conducted in SAS 9.4 and R 4.0.3.

## RESULTS

Civil registration of deaths covered 505 million people in the eight states and seven cities included in our analyses (Table 1) or 37% of India’s estimated population of 1360 million in 2019. During the first viral wave in 2020, the proportion of excess deaths compared to earlier years of registration in the subset of five included states ranged from 63% in Andhra Pradesh to 6% in Kerala (total excess 218,000 deaths; median 22%). The number of excess deaths during the first viral wave in seven cities ranged from an improbable 95% in Ahmedabad to 21% in Kolkata (total excess 82,000 deaths; median 41%).

**Table 1:**
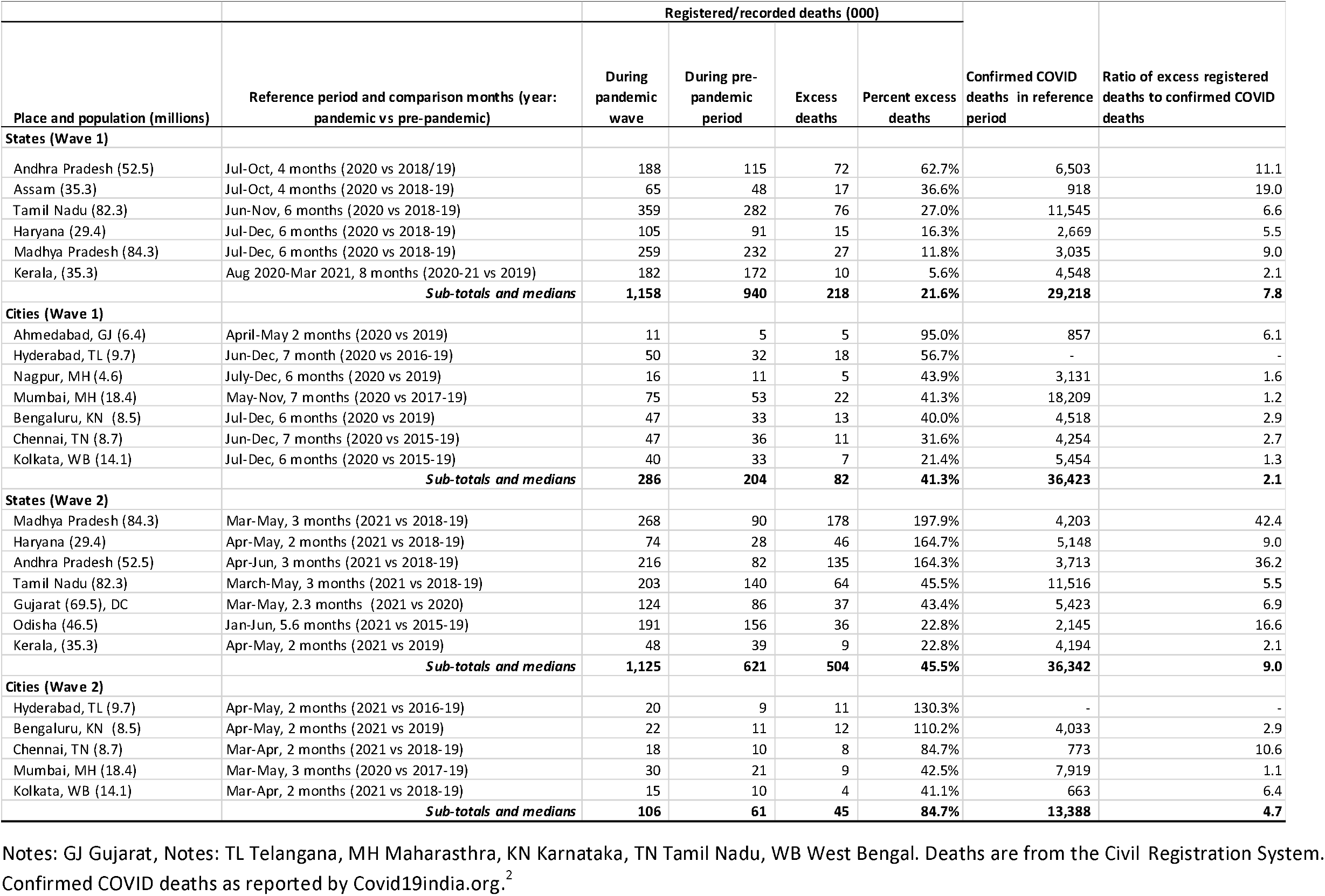
Excess deaths during COVID-19 pandemic months in 2020 and 2021, for selected states and cities in India.

During the shorter duration second viral wave of 2021, the comparable proportions for seven states ranged from 198% in Madhya Pradesh to 0% in Kerala (total excess of 495,000 deaths; median 46%). The comparable proportions for five cities was a total excess of 45,000 deaths; median 85%. The median ratio of excess to confirmed COVID deaths ranged from 8 to 9 in the two viral waves for states, and between 3 and 5 for cities.

For the subset of five states (Madhya Pradesh, Andhra Pradesh, Tamil Nadu, Kerala and Haryana) and five cities with ten or more months of observations (including in the non-pandemic months between the two viral waves), the combined excess as a percentage of expected deaths based on the UN demographic estimates was 32% for states and 37% for cities (Table 2). Results using the UN baseline estimate of deaths were similar (Table S4 in Supplementary Appendix). Total excess deaths were 0.63 million for the five states, which reported about one in five of national official COVID deaths.^2^ This implies, crudely, about 3.2 million deaths nationally in both viral waves, assuming other states have also have five times more excess deaths.

**Table 2:** Summary estimates of excess deaths in India by state and nationally for settings with 10 or more months of data.

| Location | Reference period | Months | UN-estimated deaths in reference period (000s) | Excess deaths | Excess as percentage of UN estimated deaths |
| --- | --- | --- | --- | --- | --- |
| **States** |  |  |  |  |  |
| Andhra Pradesh | July 2020-June 2021 | 12 | 411 | 207 | 50.4% |
| Haryana | July 2020-May 2021 | 11 | 191 | 61 | 31.9% |
| Madhya Pradesh | July 2020-May 2021 | 11 | 645 | 205 | 31.8% |
| Tamil Nadu | July 2020-June 2021 | 12 | 612 | 140 | 22.8% |
| Kerala | Aug 2020-May 2021 | 10 | 233 | 19 | 8.0% |
| **Sub-total and medians** |  |  | **2092** | **632** | **31.8%** |
| **Cities** |  |  |  |  |  |
| Hyderabad | June 2020-May 2021 | 12 | 52 | 30 | 57.5% |
| Bengaluru | Jul 2020-May 2021 | 11 | 48 | 25 | 52.8% |
| Chennai | June 2020-April 2021 | 11 | 53 | 20 | 36.6% |
| Mumbai | May 2020-May 2021 | 13 | 105 | 31 | 29.4% |
| Kolkata | July 2020- April 2021 | 10 | 75 | 11 | 15.2% |
| **Sub-total and medians** |  |  | **7333** | **117** | **36.6%** |
| **Facility-based deaths, national** |  |  |  |  |  |
|  | July-Dec 2020 | 6 | 1295 | 180 | 13.9% |
|  | Apr-May 2021 | 2 | 375 | 450 | 120.2% |
|  | July 2020-May 2021 | 11 | **2301** | **630** | **27.4%** |
| **Survey-based estimates, national** |  |  |  |  |  |
|  |  |  |  | <u>Lower/upper*</u> | <u>Lower/upper*</u> |
|  | June 1 2020-Jan 31 2021 | 8 | 6713 | 595/658 | 8.9%/9.8% |
|  | April 1-June 27 2021 | 3 | 2411 | 2515/2779 | 104.3%/115.3% |
|  | June 1 2020-June 27, 2021 | 13 | **10,742** | **3110/3437** | **29.0%/32.0%** |

Notes: Facility-based deaths from the Ministry of Health and Family Welfare's Health Management Information System, UN United Nations Population Division data from World Population Prospects, adjusted to Indian states/cities. \*Lower and upper estimates are based on the polling margin of error or +/- 5%

We observed a similar excess in facility-based deaths covering a non-representative sample of 0.2 million facilities in the country (Figure 1). From July 2020 to May 2021, excess deaths were 27% (total excess 0.63 million deaths) out of 2.32 million expected deaths for the 11 months (Table 2). Much of this excess (0.45 million) occurred in April-May 2021, reaching 120% versus earlier years. Excess death proportions varied across states from April-May 2021, with Gujarat reporting a 230% excess and Kerala a 37% excess. The major increase in April-May 2021 was in deaths of unknown cause, consistent with COVID effects (data not shown). The government facility deaths represent about a quarter of all deaths in India, but slightly under-representing urban areas where COVID deaths have been more commonly reported.^2^ Extrapolating facility data nationally over 12 months yields 2.7 million excess deaths from July 2020 to June 2021.

**Figure 1.**
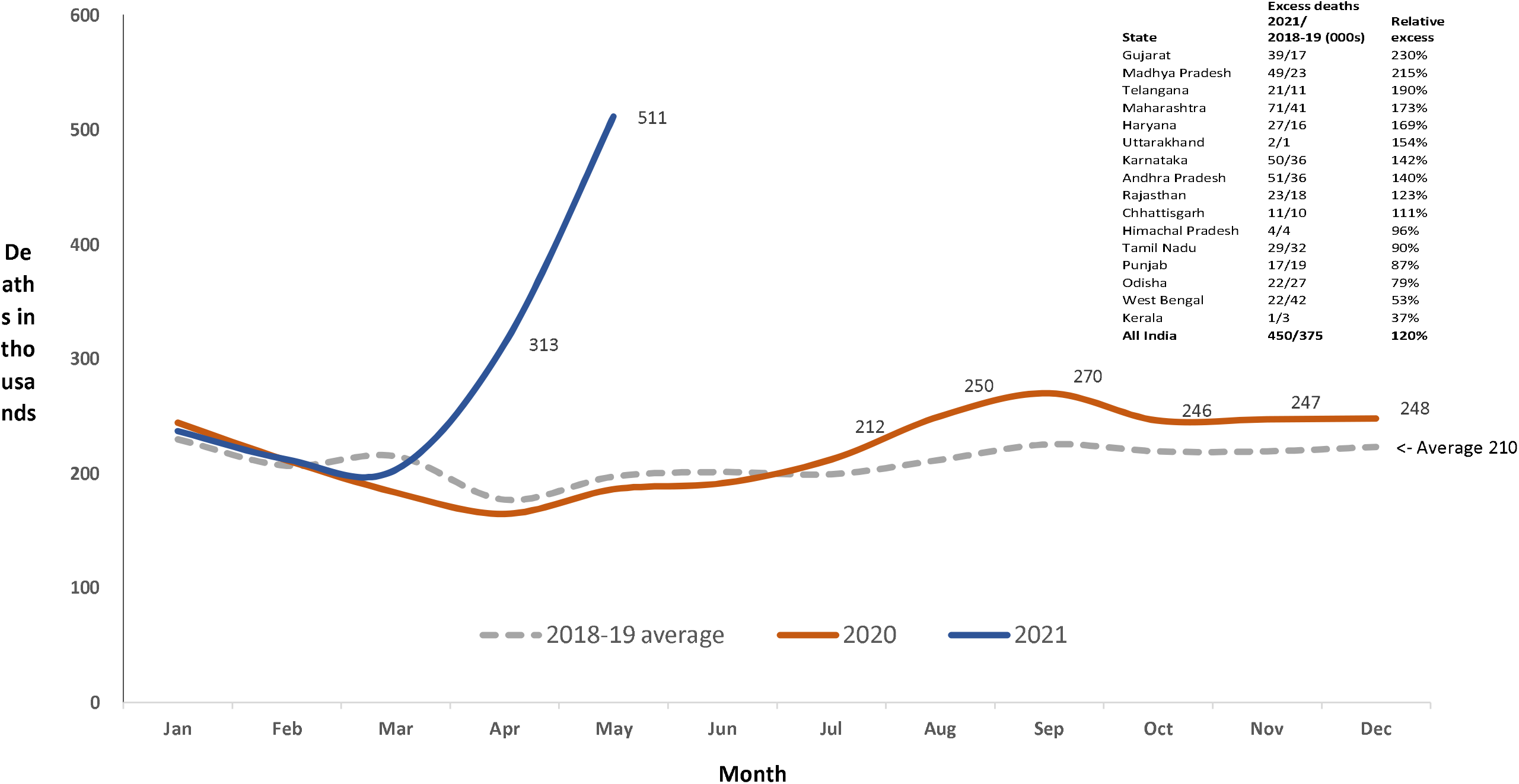
Reported deaths from all causes in India’s Ministry of Health and Family Welfare Management Information System covering 0.2 million health facilities nationally, 2020 and 2021 versus average of 2018-19, by month. The HMIS covers about 0.2 million health facilities, mostly public but some private health facilities in all states, but is not representative of the population. It reported a total of about 2.5 million deaths for the average of 2018-19, which is about 53% of the expected total of 4.7 million deaths occur in facilities.^8^ The excess percentage from July-Dec 2020 and for April-May 2021 are shown in Table 2. The inset shows results for selected states and nationally for the April-May 2021 versus 2018-19 average for the same months.

The Omnibus survey response rate was over 55%; 137,289 respondents in all states and union territories were interviewed from March 2020 to July 2021. Figure 2 displays the adjusted daily percentage of adults aged 18 years or older surveyed nationally who reported a COVID death daily (based on a smoothed 7-day rolling average of polling results) compared to reported COVID deaths as well as to the expected daily percentage for all-cause deaths based on 2019 death totals. The 10-day lagged polling daily death totals correlated strongly with the daily confirmed COVID death totals: cross-correlation 0.89, p<0.0001. There was a sharp increase in adults reporting a COVID death from April 19 to June 27, 2021, reaching peaks close to 6%, or an average of 3.5% over the 2.3 months. A smaller peak occurred over 10 days from September 24 to October 4, 2020. The 2011 census defined average household size as 4.3, which would mean about 316.3 million households nationally. Dividing this into the 10.09 million deaths in 2019 yields approximately 3.2% of households expected to report a death (irrespective of cause) per year.

**Figure 2.**
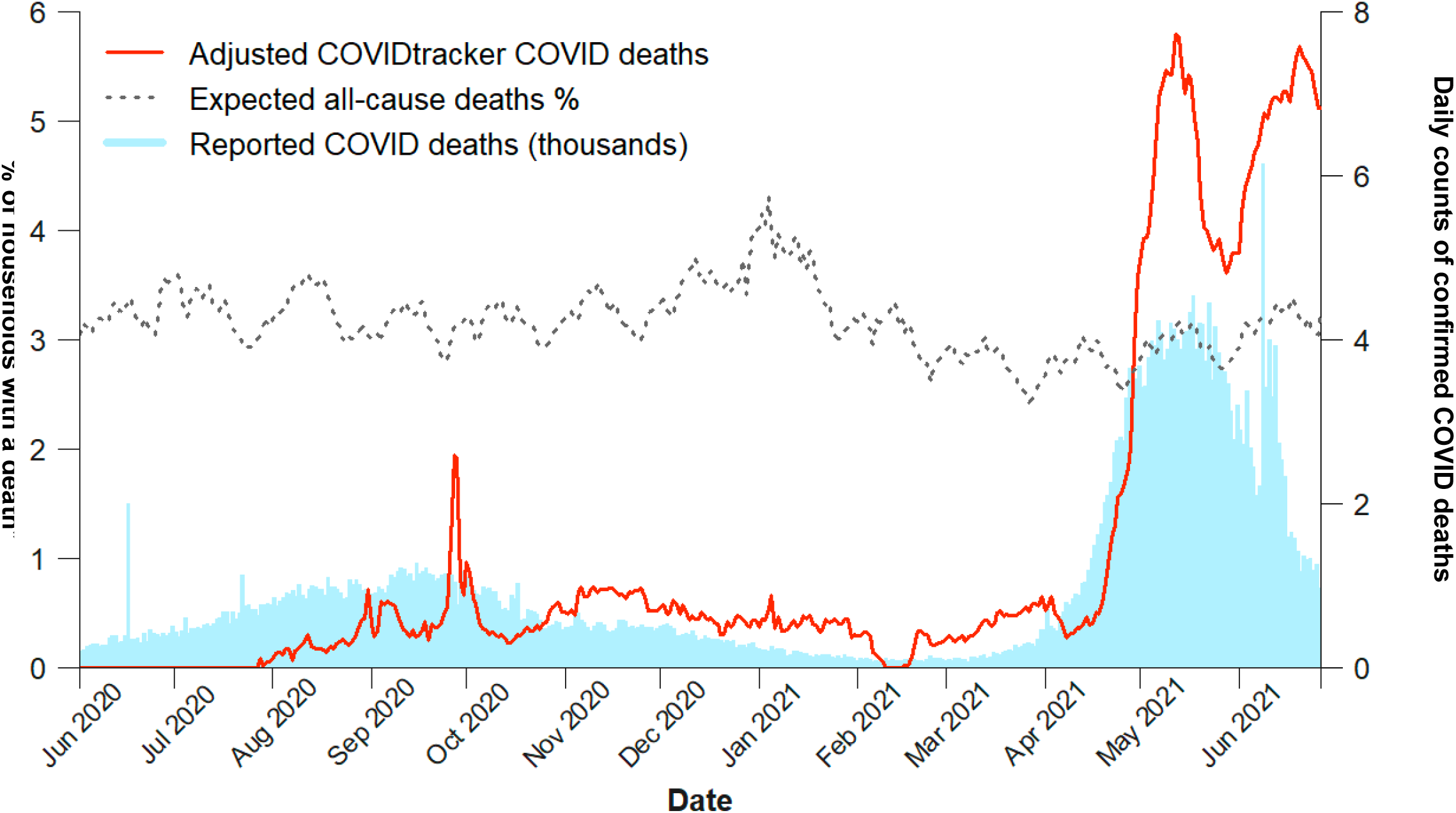
Adjusted percentages of adults reporting daily death in household, expected percentage in 2019 and daily confirmed COVID deaths in India, June 2020-June 2021. COVIDtracker deaths (red line, left vertical scale) represent COVID deaths reported daily (smoothed for rolling 7-day averages) at age 35 or older, adjusted for possible over-reporting (see text). Expected all-cause deaths (grey line, left vertical scale) per year of 3.16% applies the UN estimated total deaths in 2021 (10.0 million) to the number of households in India (316.3 million, based on the average household size of 4.3 in the 2011 census). The daily variation in this percentage applies the variation observed among 480,000 deaths in the Million Death Study. Confirmed COVID deaths (blue bars, right vertical scale) are daily reports from Covid19india.org.^2^

The April-June 2021 period showed an adjusted proportion of adults reporting COVID deaths (35 years or older) of 3.5%, which exceeded the expected all-cause proportion of 3.2% (absolute excess of 0.3%, lower and upper excess of 104% to 115%; Table 2). This yields a lower and upper estimate of 3.1 to 3.4 million excess deaths from June 1, 2020, to June 27, 2021 (Table 2). The excess deaths represent approximately 29-32% of total expected deaths in India during this period. Of these deaths, the vast majority occurred from April 1 to June 27, 2021 (2.5 to 2.8 million excess). Sensitivity analyses assigning the subtraction value for non-household reporting of COVID deaths at 0.29% yielded 3.9-4.3 million deaths and assigning it as 0.88% yielded 2.3-2.5 million deaths.

## DISCUSSION

Based on nationally representative survey data, excess deaths in India, nearly all from COVID, may have been 3.1 to 3.4 million or 29-32% of total deaths during the pandemic period of June 1, 2020, to June 27, 2021. These proportions of excess deaths are consistent with CRS estimates of 32% for selected states and 37% for selected cities and consistent with the 27% excess deaths in national government health facilities over the same time period. The range of estimates across three independent in the study data sources is between 2.7 and 3.4 million. Indeed, the COVID pandemic peaks in May-June 2021 doubled the total death rate from all conditions.

Even the lowest estimate of 2.7 million deaths and the more credible lower estimate of 3.1 million deaths represent a national COVID death rate per million population ranging from about 2000 to 2200, or approximately 6.8 to 7.9 times the reported rate of 290.^1^ This would put India’s death rate per million in the range reported in Latin America (around 2000/million), where registration of deaths is far more complete.^18^ If our findings are confirmed, this will require substantial upward revision of WHO’s global excess mortality which currently estimates three million deaths (1.2 million more than confirmed COVID deaths) in 2020 just in the Americas and European region.^9^ Indeed, our study would double the confirmed COVID deaths worldwide from January 1 to July 10, 2021, which currently stands at two million, of which 0.25 million are in India.

Definitive quantification of excess mortality can be expected once the Registrar General of India re-launches its SRS to cover all deaths occurring in 2020 and 2021. The SRS covers nearly 9,000 randomly selected villages and urban blocks to provide age- and sex-specific death rates, which will allow comparison to baseline death rates in 2018-19.^8^ The SRS is the largest, most reliable framework in place to ascertain rural death rates in India. The Centre for Monitoring Indian Economy national household survey also reports a large excess of deaths,^19^ but has uncertain demographic parameters (which is one reason why we excluded it from our analyses). Most urban municipalities in India maintain daily records of the death registrations or requests for death certificates.^5^ Release of weekly totals of such data from 2018 onwards would be useful. These data are essential not only for a complete accounting of the large death total from COVID but also to help identify geographic pockets and demographic groups (such as women or scheduled castes and other socially disadvantaged groups) with excess mortality, likely representing increased community transmission. Indeed, the states with the lowest levels of death registration reported the lowest confirmed case rates, suggesting that information gaps in mortality may also apply to infectious disease surveillance (Figure S1 in Supplementary Appendix). Moreover, disaggregated, local mortality information could help target expansion of SARS-CoV-2 vaccines, helping to minimize the size of the inevitable third viral wave projected to hit India in the fall of 2021.^13^

In the longer term, India must expand and improve its death registration and medical certification system including timelier reporting.^20^ India still does not register about three million deaths annually and does not certify the medical cause in about eight million. These gaps are not equal, being largest for the poorest states in central India and much greater for women than for men (Figure S2 and Table S5 in Supplementary Appendix).

The strengths of our study are its nationally representative and distributed sampling for the survey, use of three independent data sources, and robust metrics that document excess deaths versus earlier years or expected demographic data. Moreover, our methods are reproducible over time and avoid the limitations of model-based estimates. Nonetheless, we faced some limitations. The use of excess mortality has limitations as some causes, notably road traffic accidents and other injuries, might have fallen, particularly during COVID lockdowns. However, these injuries constitute less than 10% of all deaths from 2004-14.^4,21^ There might also be an excess of some deaths from neglected health service, and there are reports, for example, that maternal mortality has risen during the pandemic months.^22^ However, changes in causes of death aside from COVID are likely small compared to the sharp increases in COVID deaths, particularly during the second viral wave. The delays in death registration or a backlog of deaths reported suddenly might create an artifact of excess deaths. However, in the case of Andhra Pradesh, 98% of deaths registered in May 2021 took place within the previous 30 days, not earlier time periods. Survey data might represent over-reporting, as the questions asked about deaths beyond the households. However, since the introduction was focused on flu-like symptoms in the respondent’s family or daily contacts, the extent of such over-reporting is likely small. Moreover, we corrected for over-reporting, albeit imperfectly, by subtracting reported rates in non-pandemic weeks and tested this assumption in sensitivity analyses, which suggested that a lowest value of 2.3 million COVID deaths over the study year. New surveys under way will more directly ascertain household composition prior to the pandemic and subsequent deaths from any cause, and will help to resolve these uncertainties.

In sum, our study finds that Indian COVID deaths are substantially greater than estimated from official reports. Mortality data will be central to help assess and control the inevitable third viral wave expected in India.

## Data Availability

The data collected for this study will be made available to others online.

Supported by Canadian Institutes of Health Research (FDN 154277 and 20928). The Canada Research Chairs Program funds PJ.

We thank Rukmini S, Srinivasan Ramani, Vignesh Radhakhrishnan, Anurabh Saikia, Mariyam Alavi, Saurav Das, Pratap Vardhan and Dhanya Rajendran who were crucial in filing petitions to access the various data sources.

