## Supplementary Appendix for "Excess mortality in India from June 2020 to June 2021 during the COVID pandemic: death registration, health facility deaths, and survey data"

### Table S1. Input data used to create Table 1

| Year / No. of deaths | 2015 | 2016 | 2017 | 2018 | 2019 | 2020 | 2021 | Pandemic period (months) | | | Total deaths |
|---|---|---|---|---|---|---|---|---|---|---|---|
| Andhra Pradesh State (GitHub-DDL, GitHub-local mortality) | | | | | | | | | | | |
| 1 | | | | 33,637 | 32,302 | 31,989 | 37,963 | | | | |
| 2 | | | | 28,788 | 31,170 | 26,802 | 31,920 | | Andhra Pradesh | | |
| 3 | | | | 27,341 | 27,972 | 25,413 | 33,367 | 1st Wave: | Jul-OCT 2020 | 4m | 187,675 |
| 4 | | | | 25,979 | 26,290 | 22,362 | 39,102 | 2nd Wave | Apr-Jun 2021 | 3m | 216,439 |
| 5 | | | | 24,357 | 29,928 | 28,066 | 134,041 | | | | |
| 6 | | | | 27,006 | 30,201 | 28,440 | 43,296 | | | | |
| 7 | | | | 26,458 | 28,465 | 34,757 | | | | | |
| 8 | | | | 27,929 | 28,667 | 54,940 | | | | | |
| 9 | | | | 26,626 | 30,447 | 55,874 | | | | | |
| 10 | | | | 28,478 | 33,631 | 42,104 | | | | | |
| 11 | | | | 26,785 | 33,201 | 38,665 | | | | | |
| 12 | | | | 29,891 | 31,140 | 39,495 | | | | | |
| Assam State (GitHub-local mortality) | | | | | | | | | | | |
| 1 | | | | 12,204 | 15,680 | 18,556 | | | Assam | | |
| 2 | | | | 12,193 | 14,806 | 17,971 | | 1st Wave: | Jul-Oct 2020 | 4m | 65,183 |
| 3 | | | | 12,155 | 14,266 | 16,975 | | 2nd Wave | Apr-Jun 2021 | 2m | NA |
| 4 | | | | 11,826 | 13,147 | 9,003 | | | | | |
| 5 | | | | 11,761 | 12,169 | 8,655 | | | | | |
| 6 | | | | 11,656 | 11,303 | 8,305 | | | | | |
| 7 | | | | 11,456 | 11,065 | 9,118 | | | | | |
| 8 | | | | 11,378 | 10,925 | 16,887 | | | | | |
| 9 | | | | 11,593 | 13,177 | 18,364 | | | | | |
| 10 | | | | 11,755 | 14,117 | 20,814 | | | | | |
| 11 | | | | 11,515 | 15,700 | 21,780 | | | | | |
| 12 | | | | 13,113 | 16,702 | 20,657 | | | | | |
| Madhya Pradesh State (GitHub-local mortality) | | | | | | | | | | | |
| 1 | | | | 34,451 | 39,528 | 41,281 | 44,133 | | Madhya Pradesh | | |
| 2 | | | | 29,986 | 36,005 | 34,645 | 36,535 | 1st Wave: | Jul-Dec 2020 | 6m | 259,467 |
| 3 | | | | 28,504 | 32,911 | 29,747 | 34,667 | 2nd Wave | Mar-May 2021 | 3m | 268,040 |
| 4 | | | | 26,949 | 28,599 | 24,198 | 68,535 | | | | |
| 5 | | | | 32,456 | 30,506 | 34,320 | 164,838 | | | | |
| 6 | | | | 33,818 | 38,978 | 37,399 | | | | | |
| 7 | | | | 36,523 | 38,922 | 41,303 | | | | | |
| 8 | | | | 38,230 | 44,151 | 40,213 | | | | | |
| 9 | | | | 40,180 | 43,517 | 47,315 | | | | | |
| 10 | | | | 37,766 | 37,953 | 45,208 | | | | | |
| 11 | | | | 30,349 | 39,124 | 40,283 | | | | | |
| 12 | | | | 37,960 | 39,625 | 45,145 | | | | | |
| Tamil Nadu State (GitHub-local mortality) | | | | | | | | | | | |
| 1 | | | | 55,390 | 58,405 | 58,132 | 62,273 | | Tamil Nadu | | |
| 2 | | | | 47,231 | 46,560 | 48,893 | 52,845 | 1st Wave: | Jun-Nov 2020 | 6m | 358,546 |
| 3 | | | | 46,703 | 47,772 | 45,987 | 50,959 | 2nd Wave | March-May 20[illegible] | 3m | 203,307 |
| 4 | | | | 43,586 | 45,913 | 41,482 | 58,775 | | | | |
| 5 | | | | 43,928 | 51,639 | 50,834 | 93,573 | | | | |
| 6 | | | | 43,222 | 48,868 | 50,687 | | | | | |
| 7 | | | | 42,633 | 45,762 | 60,052 | | | | | |
| 8 | | | | 41,051 | 47,385 | 70,033 | | | | | |
| 9 | | | | 40,484 | 48,621 | 65,447 | | | | | |
| 10 | | | | 46,337 | 53,028 | 60,096 | | | | | |
| 11 | | | | 51,602 | 55,658 | 52,231 | | | | | |

| | | | | | | | | | | | |
|---|---|---|---|---|---|---|---|---|---|---|---|
| 12 | | | | 34,025 | 38,610 | 40,417 | | | | | |
| Kerala state (GitHub-DDL) | | | | | | | | | | | |
| 1 | | | 20,326 | 22,081 | 23,676 | 21,357 | 23,328 | | Kerala | | |
| 2 | | | 18,128 | 19,354 | 19,892 | 19,714 | 21,152 | 1st Wave: | Aug 2020-Mar | 8m | 181,754 |
| 3 | | | 19,013 | 19,843 | 20,640 | 19,581 | 20,706 | 2nd Wave | Apr-Jun 2021 | 3m | 54,094 |
| 4 | | | 19,014 | 18,825 | 19,983 | 17,212 | 20,229 | | | | |
| 5 | | | 20,053 | 19,370 | 20,514 | 17,680 | 27,957 | | | | |
| 6 | | | 22,597 | 20,435 | 20,509 | 19,346 | 5,908 | | | | |
| 7 | | | 26,302 | 23,372 | 24,567 | 20,963 | . | | | | |
| 8 | | | 23,101 | 25,683 | 25,475 | 23,394 | . | | | | |
| 9 | | | 21,513 | 22,844 | 24,318 | 22,704 | . | | | | |
| 10 | | | 20,681 | 21,075 | 22,005 | 24,949 | . | | | | |
| 11 | | | 20,083 | 21,031 | 21,626 | 22,115 | . | | | | |
| 12 | | | 21,292 | 21,681 | 20,945 | 23,406 | . | | | | |
| Gujarat state (Media published Death Certificate issues and CR data) | | | | | | | | | | | |
| 3 | | | | | | | 26,026 | 2nd Wave | Mar-May, 2.3 n | 2.3m | 123,873 |
| 4 | | | | | 86,410 | | 57,796 | | | | |
| 5* | | | | | CR Report | | 40,051 | * Up to May 10th | | | |
| Haryana state | | | | | | | | | | | |
| 1 | | | | 17,818 | 17,638 | 19,066 | 17,858 | | Haryana | | |
| 2 | | | | 14,614 | 15,268 | 15,727 | 14,908 | 1st wave | Jul-Dec 2020 | 6 | 105,417 |
| 3 | | | | 14,567 | 15,316 | 14,787 | 14,908 | 2nd Wave | Apr-May 2021 | 2 | 74,384 |
| 4 | | | | 13,580 | 13,658 | 12,965 | 28,276 | | | | |
| 5 | | | | 14,335 | 14,630 | 15,445 | 46,108 | | | | |
| 6 | | | | 14,772 | 14,946 | 15,496 | | | | | |
| 7 | | | | 13,115 | 13,486 | 15,590 | | | | | |
| 8 | | | | 13,648 | 14,871 | 15,581 | | | | | |
| 9 | | | | 14,079 | 14,861 | 17,253 | | | | | |
| 10 | | | | 15,564 | 15,003 | 16,611 | | | | | |
| 11 | | | | 15,941 | 15,527 | 20,914 | | | | | |
| 12 | | | | 16,601 | 18,591 | 19,468 | | | | | |
| Ahmedabad City (News paper reporting) | | | | | | | | | | | |
| 4 | | | | | 2,784 | 3,558 | | 1st Wave: | Apr-May 2020 | 2m | 10,708 |
| 5 | | | | | 2,706 | 7,150 | | | | | |
| Bangalore (Urban) (GitHub-DDL) | | | | | | | | | | | |
| 1 | | | . | . | 5,168 | 5,983 | 6,012 | | Bangalore (Urban) | | |
| 2 | | | . | . | 5,766 | 5,454 | 5,538 | 1st Wave: | Jul-Dec 2020 | 6m | 46,611 |
| 3 | | | . | . | 5,400 | 4,716 | 6,216 | 2nd Wave | Apr-May 2021 | 2m | 22,498 |
| 4 | | | . | . | 4,806 | 3,327 | 6,249 | | | | |
| 5 | | | . | . | 5,899 | 4,469 | 16,249 | | | | |
| 6 | | | . | . | 4,687 | 4,881 | . | | | | |
| 7 | | | . | . | 5,278 | 6,477 | . | | | | |
| 8 | | | . | . | 5,481 | 9,340 | . | | | | |
| 9 | | | . | . | 5,411 | 8,710 | . | | | | |
| 10 | | | . | . | 5,597 | 8,413 | . | | | | |
| 11 | | | . | . | 5,982 | 7,342 | . | | | | |
| 12 | | | . | . | 5,544 | 6,329 | . | | | | |
| Chennai City (GitHub-local mortality) | | | | | | | | | | | |
| 1 | 5,543 | 5,644 | 5,833 | 5,837 | 6,228 | 5,915 | 6,195 | | Chennai City | | |
| 2 | 4,753 | 4,528 | 4,877 | 5,132 | 5,114 | 5,046 | 5,218 | 1st wave | Jun-Dec 2020 (6n | 7m | 47,277 |
| 3 | 4,959 | 4,633 | 4,925 | 5,107 | 5,286 | 4,961 | 6,038 | 2nd wave | Mar-Apr 2021 | 2m | 17,914 |
| 4 | 4,505 | 4,611 | 4,735 | 4,772 | 4,964 | 4,149 | 11,876 | | | | |
| 5 | 5,072 | 5,102 | 8,142 | 5,214 | 5,758 | 6,650 | 5,815 | | | | |
| 6 | 4,698 | 4,690 | 4,890 | 5,065 | 6,071 | 7,747 | - | | | | |
| 7 | 4,499 | 4,717 | 5,156 | 5,003 | 5,121 | 6,368 | - | | | | |

| | | | | | | | | | | | |
|---|---|---|---|---|---|---|---|---|---|---|---|
| 8 | 4,484 | 4,712 | 5,259 | 4,814 | 5,351 | 6,805 | - | | | | |
| 9 | 4,460 | 4,609 | 5,728 | 4,828 | 5,457 | 7,087 | - | | | | |
| 10 | 5,019 | 4,572 | 5,603 | 5,762 | 6,147 | 6,714 | - | | | | |
| 11 | 6,064 | 4,756 | 5,470 | 5,770 | 5,978 | 6,115 | - | | | | |
| 12 | 5,842 | 4,610 | 4,691 | 4,802 | 4,885 | 6,440 | - | | | | |
| Hyderabad (GitHub-local mortality) | | | | | | | | | | | |
| 1 | | 4,033 | 4,455 | 4,841 | 5,470 | 5,763 | 5,879 | | Hyderabad City | | |
| 2 | | 3,319 | 3,613 | 4,354 | 4,628 | 5,199 | 4,749 | 1st Wave: | Jun-Dec 2020 | 7m | 50,337 |
| 3 | | 3,664 | 4,035 | 4,277 | 4,851 | 4,894 | 5,090 | 2nd Wave | Apr-May 2021 | 2m | 20,323 |
| 4 | | 3,979 | 4,016 | 4,002 | 4,708 | 3,967 | 9,465 | | | | |
| 5 | | 3,844 | 4,526 | 4,222 | 6,000 | 5,061 | 10,858 | | | | |
| 6 | | 3,567 | 3,669 | 3,746 | 4,707 | 7,011 | | | | | |
| 7 | | 4,152 | 3,954 | 4,103 | 4,808 | 10,423 | | | | | |
| 8 | | 4,525 | 4,220 | 4,271 | 5,620 | 7,974 | | | | | |
| 9 | | 3,961 | 4,525 | 4,615 | 6,048 | 7,013 | | | | | |
| 10 | | 4,255 | 4,650 | 4,925 | 6,117 | 6,212 | | | | | |
| 11 | | 4,087 | 4,427 | 4,700 | 5,466 | 5,708 | | | | | |
| 12 | | 4,121 | 4,603 | 4,952 | 5,689 | 5,996 | | | | | |
| Kolkata City (GitHub-local mortality) | | | | | | | | | | | |
| 1 | 6,441 | 6,598 | 7,103 | 8,506 | 7,918 | 7,587 | 6,293 | | Kolkata City | | |
| 2 | 5,598 | 4,889 | 5,646 | 5,934 | 6,421 | 6,115 | 4,776 | 1st wave | Jul-Dec 2020 (6m | 6m | 40,482 |
| 3 | 5,279 | 4,923 | 5,434 | 5,346 | 6,078 | 5,377 | 4,374 | 2nd wave | Mar-Apr 2021 | 2m | 14,630 |
| 4 | 4,582 | 4,978 | 5,246 | 4,861 | 5,128 | 4,830 | 10,256.29 | | | | |
| 5 | 4,890 | 4,669 | 5,476 | 4,904 | 5,314 | 5,375 | 6,615.86 | | | | |
| 6 | 4,421 | 4,783 | 4,730 | 5,066 | 5,395 | 5,007 | - | | | | |
| 7 | 4,754 | 5,391 | 5,549 | 4,915 | 5,329 | 6,590 | - | | | | |
| 8 | 5,274 | 5,610 | 6,275 | 5,532 | 5,056 | 6,521 | - | | | | |
| 9 | 5,389 | 5,257 | 5,704 | 5,612 | 5,092 | 6,050 | - | | | | |
| 10 | 5,194 | 5,619 | 5,905 | 5,804 | 5,836 | 6,986 | - | | | | |
| 11 | 5,170 | 5,924 | 6,337 | 5,685 | 5,805 | 7,238 | - | | | | |
| 12 | 5,739 | 5,665 | 5,747 | 6,012 | 5,502 | 7,096 | - | | | | |
| Nagpur City (GitHub-local mortality) | | | | | | | | | | | |
| 3 | | | | | | 1,415 | 3,168 | | | | |
| 4 | | | | | 1,583 | 1,311 | | | Nagpur | | |
| 5 | | | | | 1,900 | 1,624 | | 1st wave | July-Dec2020 | 6m | 15,935 |
| 6 | | | | | 1,678 | 1,512 | | 2nd Wave | Mar-May 2021 | 3m | |
| 7 | | | | | 1,590 | 1,808 | | | | | |
| 8 | | | | | 1,787 | 3,385 | | | | | |
| 9 | | | | | 1,958 | 4,096 | | | | | |
| 10 | | | | | 1,801 | 2,387 | | | | | |
| 11 | | | | | 1,775 | 2,325 | | | | | |
| 12 | | | | | 2,166 | 1,934 | | | | | |
| Mumbai City (GitHub-local mortality) | | | | | | | | | | | |
| 1 | | | 8,004 | 8,306 | 8,324 | 8,397 | 7,732 | | Mumbai city | | |
| 2 | | | 7,158 | 7,305 | 7,797 | 7,116 | 7,131 | 1st wave | May-Nov2020 | 7m | 74,811 |
| 3 | | | 7,810 | 7,436 | 7,155 | 5,703 | 8,302 | 2nd Wave | Mar-June 2021 | 4m | 30,300 |
| 4 | | | 6,234 | 6,719 | 6,752 | 5,537 | 14,484 | | | | |
| 5 | | | 6,960 | 7,407 | 7,335 | 9,161 | 7,514 | | | | |
| 6 | | | 7,068 | 6,874 | 6,732 | 15,756 | . | | | | |
| 7 | | | 7,675 | 7,336 | 7,931 | 11,770 | . | | | | |
| 8 | | | 7,247 | 7,372 | 8,164 | 10,215 | . | | | | |
| 9 | | | 8,250 | 7,231 | 7,953 | 10,061 | . | | | | |
| 10 | | | 7,700 | 8,755 | 7,390 | 9,835 | . | | | | |
| 11 | | | 7,943 | 7,235 | 8,320 | 8,013 | . | | | | |
| 12 | | | 6,988 | 6,876 | 7,370 | 7,834 | | | | | |

National (HMIS, Ministry of Health and Family Welfare, GoI)

| | | | | | | | | | | |
|---|---|---|---|---|---|---|---|---|---|---|
| 1 | | | 221,543 | 238,079 | 244,345 | 234,907 | | National (HMIS) | | |
| 2 | | | 204,119 | 209,475 | 211,623 | 212,211 | 1st wave | Jul-Dec 2020 | 6m | 1,474,997 |
| 3 | | | 192,123 | 198,159 | 183,436 | 203,316 | 2nd Wave | Apr-May 2021 | 2m | 825,074 |
| 4 | | | 167,999 | 186,274 | 164,831 | 311,688 | | | | |
| 5 | | | 188,632 | 206,479 | 186,395 | 513,386 | | | | |
| 6 | | | 191,197 | 211,723 | 191,544 | | | | | |
| 7 | | | 191,412 | 207,594 | 212,556 | | | | | |
| 8 | | | 197,917 | 226,277 | 250,096 | | | | | |
| 9 | | | 210,200 | 232,270 | 270,371 | | | | | |
| 10 | | | 214,351 | 224,700 | 246,526 | | | | | |
| 11 | | | 213,179 | 225,872 | 247,578 | | | | | |
| 12 | | | 215,885 | 230,308 | 247,870 | | | | | |
| | | | 2,408,557 | 2,597,210 | 2,657,171 | 1,475,508 | | | | |

Uttar Pradesh state (GitHub-DDL)

| | | | | | | | | | | |
|---|---|---|---|---|---|---|---|---|---|---|
| 1 | . | . | | 68,962 | 75,773 | 103,048 | | Uttar Pradesh | | |
| 2 | . | . | | 69,809 | 50,576 | 98,047 | 1st Wave: | Jul-Dec 2020 | 6m | 510,175 |
| 3 | . | . | | 63,962 | 41,683 | 70,797 | 2nd Wave | Apr-21 | 1m | 61,986 |
| 4 | . | . | | 56,583 | 20,715 | 61,986 | | | | |
| 5 | . | . | | 55,338 | 33,543 | . | | | | |
| 6 | . | . | | 62,657 | 59,020 | . | | | | |
| 7 | . | . | | 63,604 | 68,382 | . | | | | |
| 8 | . | . | | 67,811 | 78,658 | . | | | | |
| 9 | . | . | | 69,589 | 92,947 | . | | | | |
| 10 | . | . | | 60,004 | 90,023 | . | | | | |
| 11 | . | . | | 66,740 | 80,981 | . | | | | |
| 12 | . | . | | 66,324 | 99,184 | . | | | | |

Bihar state (GitHub-DDL)

| | | | | | | | | | | |
|---|---|---|---|---|---|---|---|---|---|---|
| 1 | . | | 7,289 | 28,975 | 38,238 | 45,515 | | Bihar | | |
| 2 | . | | 7,872 | 27,620 | 34,015 | 38,981 | 1st Wave: | Jul-Dec 2020 | 6m | 216,080 |
| 3 | . | | 8,196 | 28,452 | 26,682 | 29,774 | 2nd Wave | Apr-May 2021 | 2m | 101,476 |
| 4 | . | | 9,635 | 23,537 | 16,865 | 33,159 | | | | |
| 5 | . | | 12,495 | 24,640 | 22,893 | 68,317 | | | | |
| 6 | . | | 13,898 | 30,440 | 32,656 | . | | | | |
| 7 | . | | 15,926 | 31,533 | 30,534 | . | | | | |
| 8 | . | | 17,933 | 34,326 | 36,355 | . | | | | |
| 9 | . | | 20,480 | 27,897 | 39,124 | . | | | | |
| 10 | . | | 22,413 | 29,168 | 31,860 | . | | | | |
| 11 | . | | 22,317 | 32,088 | 30,299 | . | | | | |
| 12 | . | | 24,467 | 32,598 | 47,908 | . | | | | |

Rajasthan (GitHub-DDL)

| | | | | | | | | | | |
|---|---|---|---|---|---|---|---|---|---|---|
| 1 | | | 20,798 | 20,239 | 21,954 | 19,622 | | Rajasthan | | |
| 2 | | | 18,301 | 18,089 | 18,056 | 14,860 | 1st wave | Jul-Dec 2020 | 6m | 117,039 |
| 3 | | | 18,921 | 17,784 | 16,378 | 16,084 | 2nd Wave | Apr-May 2021 | 2m | 75,295 |
| 4 | | | 18,018 | 16,651 | 17,596 | 26,251 | | | | |
| 5 | | | 19,446 | 17,603 | 20,582 | 49,044 | | | | |
| 6 | | | 17,048 | 19,766 | 17,959 | | | | | |
| 7 | | | 15,640 | 15,656 | 17,151 | | | | | |
| 8 | | | 16,023 | 17,569 | 17,918 | | | | | |
| 9 | | | 15,812 | 19,291 | 18,719 | | | | | |
| 10 | | | 17,226 | 17,924 | 18,405 | | | | | |
| 11 | | | 17,489 | 17,794 | 21,265 | | | | | |
| 12 | | | 21,648 | 21,448 | 23,581 | | | | | |

**Table S2 – Comparison of Cvoter sample characteristics with 2011 Indian Census data or National Sample Survey**

| Characteristics | Census 2011 | Valid Sample Percent | Deviation |
|---|---|---|---|
| **Population** | | | |
| Andhra Pradesh | 4.3 | 4.3 | 0 |
| Assam | 2.3 | 2.3 | 0 |
| Bihar | 7.6 | 7.6 | 0 |
| Chhattisgarh | 2.2 | 2.1 | 0.1 |
| Delhi | 1.5 | 1.5 | 0 |
| Goa | 0.1 | 0.1 | 0 |
| Gujarat4.9 | 4.9 | 0 | |
| Haryana | 1.9 | 1.9 | 0 |
| Himachal Pradesh | 0.6 | 0.6 | 0 |
| Jammu & Kashmir | 0.9 | 0.9 | 0 |
| Jharkhand | 2.4 | 2.4 | 0 |
| Karnataka | 5.5 | 5.5 | 0 |
| Kerala | 2.9 | 2.9 | 0 |
| Madhya Pradesh | 5.7 | 5.7 | 0 |
| Maharashtra | 9.8 | 9.8 | 0 |
| NE | 1.1 | 1.1 | 0 |
| Orissa | 3.5 | 3.5 | 0 |
| Punjab | 2.4 | 2.4 | 0 |
| Rajasthan | 5.2 | 5.2 | 0 |
| Tamil Nadu | 6.6 | 6.6 | 0 |
| UT | 0.3 | 0.3 | 0 |
| Uttar Pradesh | 16.5 | 16.5 | 0 |
| Uttarakhand | 0.8 | 0.8 | 0 |
| West Bengal | 7.7 | 7.7 | 0 |
| Telangana | 3.3 | 3.3 | 0 |
| Total All India | 100 | 100 | |
| **Gender** | | | |
| Male | 52 | 52 | 0 |
| Female | 48 | 48 | 0 |
| Total | 100 | | |
| **Age Group** | | | |
| Fresher(Below-25) | 21.7 | 21.8 | -0.1 |
| Young(25-45) | 47.1 | 47.1 | 0 |
| Middle Aged(45-60) | 18.5 | 18.5 | 0 |
| Old(60 and above) | 12.7 | 12.6 | 0.1 |
| Total | 100 | 100 | |
| **Education** | | | |
| Lower Education | 67 | 67 | 0 |
| Middle Education | 20 | 20 | 0 |
| Higher Education | 13 | 13 | 0 |
| Total | 100 | 100 | |
| **Income** | | | |
| Lower Income Group | 50 | 50.1 | -0.1 |
| Middle Income Group | 35 | 35 | 0 |
| Higher Income Group | 15 | 14.9 | 0.1 |
| Total | 100 | 100 | |
| **Social Group** | | | |

| | | | |
|---|---|---|---|
| UCH (Upper Caste Hindus) | 23.7 | 23.8 | -0.1 |
| SC (Scheduled Caste/Dalit) | 16.2 | 16.2 | 0 |
| OBC (Other Backward Classes) | 32 | 32.1 | -0.1 |
| ST (Scheduled Tribes) | 8.2 | 8.2 | 0 |
| Muslim | 13.7 | 13.7 | 0 |
| Christians | 2.3 | 2.3 | 0 |
| Others | 1.9 | 1.9 | 0 |
| Sikhs | 1.9 | 1.8 | 0.1 |
| Total | 100 | 100 | |
| **Location** | | | |
| Urban | 30 | 30 | 0 |
| Rural | 70 | 70 | 0 |
| Total | 100 | 100 | |

**Table S3: SRS and estimated CGHR-UN death rates in major Indian states**

| India & bigger States | Death rate/1000 | | | | |
|---|---|---|---|---|---|
| | CGHR-UN 2019 | | | CGHR-UN 2018 | SRS 2018 |
| | Both Sexes | Male | Female | both sexes | both sexes |
| India | 7.3 | 7.6 | 7.0 | 7.3 | 6.2 |
| Assam | 5.9 | 6.1 | 5.7 | 6.0 | 6.4 |
| Jammu & Kashmir | 4.5 | 5.2 | 3.8 | 4.6 | 4.9 |
| Punjab | 6.4 | 6.7 | 6.0 | 6.4 | 6.6 |
| Himachal Pradesh | 6.3 | 7.2 | 5.3 | 6.3 | 6.9 |
| Odisha | 7.5 | 8.3 | 6.8 | 7.6 | 7.3 |
| Chhattisgarh | 8.4 | 9.1 | 7.7 | 8.4 | 8.0 |
| Bihar | 6.2 | 5.8 | 6.6 | 6.2 | 5.8 |
| Jharkhand | 5.8 | 5.6 | 6.1 | 5.9 | 5.4 |
| Gujarat | 6.4 | 7.0 | 5.8 | 6.4 | 5.9 |
| Kerala | 7.9 | 8.8 | 7.1 | 7.7 | 6.9 |
| Tamil Nadu | 7.5 | 8.3 | 6.7 | 7.5 | 6.5 |
| Andhra Pradesh | 7.8 | 7.9 | 7.7 | 7.8 | 6.7 |
| West Bengal | 6.7 | 6.7 | 6.6 | 6.6 | 5.6 |
| Haryana | 7.1 | 7.6 | 6.5 | 7.1 | 5.9 |
| Rajasthan Nadu | 7.4 | 8.4 | 6.3 | 7.2 | 5.9 |
| Uttar Pradesh | 8.3 | 8.5 | 8.0 | 8.2 | 6.6 |
| Maharashtra | 6.9 | 6.8 | 7.1 | 6.9 | 5.5 |
| Madhya Pradesh | 8.4 | 9.1 | 7.8 | 8.4 | 6.7 |
| Delhi | 4.2 | 4.7 | 3.7 | 4.3 | 3.3 |
| Karnataka | 9.3 | 9.3 | 9.3 | 9.1 | 6.3 |
| Uttarakhand | 9.4 | 9.3 | 9.4 | 9.3 | 6.2 |

Notes

* SRS Death rates are as published by Registrar General of India's SRS Statistical Report 2018

** CGHR (Centre for Global Health Research)-UN Death rates were calculated while applying the 2016-18 average death rates to calculate estimated deaths and then adjusting to the United Nations population Division estimated India deaths.

### Table S4: Excess deaths compared to UN baseline rates and completeness of death registration in Indian states

| | | Registered/r | Expected deaths | | | |
|---|---|---|---|---|---|---|
| Place and population (millions) | Reference period and comparison months (year: pandemic vs pre-pandemic) | During pandemic wave | UN/SRS-based Expected deaths for relevant months | Excess deaths as % of expected death | RGI reported coverage of death registration | Actual coverage of death registration |
| **States (Wave 1)** | | | | | | |
| Andhra Pradesh (52.5) | Jul-Oct, 4 months (2020 vs 2018/19) | 188 | 137 | 37.1% | 100% | 84% |
| Assam (35.3) | Jul-Oct, 4 months (2020 vs 2018-19) | 65 | 69 | | 74% | 69% |
| Tamil Nadu (82.3) | Jun-Nov, 6 months (2020 vs 2018-19) | 359 | 306 | 17.2% | 100% | 92% |
| Haryana (29.3) | Jul-Dec, 6 months (2020 vs 2018-19) | 105 | 104 | 0.9% | 100% | 87% |
| Madhya Pradesh (84.3) | Jul-Dec, 6 months (2020 vs 2018-19) | 259 | 352 | | 89% | 66% |
| Kerala, (35.3) | Aug 2020-Mar 2021, 8 months (2020-21 vs 2019) | 182 | 186 | | 100% | 92% |
| | ***Sub-totals and medians*** | **1,158** | **1,155** | **17.2%** | **85.5%** | **85.5%** |
| **Cities (Wave 1)** | | | | | | |
| Ahmedabad, GJ (6.4) | April-May 2 months (2020 vs 2019) | 11 | 6,130 | 74.7% | 100% | 90% |
| Hyderabad, TL (9.7) | Jun-Dec, 7 month (2020 vs 2016-19) | 50 | 30,148 | 67.0% | 97% | 107% |
| Nagpur, MH (4.6) | July-Dec, 6 months (2020 vs 2019) | 16 | 12,120 | 31.5% | 100% | 91% |
| Mumbai, MH (18.4) | May-No, 7 months (2020 vs 2017-19) | 75 | 56,560 | 32.3% | 100% | 94% |
| Bengaluru, KN (8.5) | Jul-Dec, 6 months (2020 vs 2019) | 47 | 25,925 | 79.8% | 100% | 128% |
| Chennai, TN (8.7) | Jun-Dec, 7 months (2020 vs 2015-19) | 47 | 34,030 | 38.9% | 100% | 106% |
| Kolkata, WB (14.1) | Jul-Dec, 6 months (2020 vs 2015-19) | 40 | 44,988 | 0.0% | 100% | 74% |
| | ***Sub-totals and medians*** | **286** | **209,901** | **38.9%** | **100.0%** | **93.6%** |
| **States (Wave 2)** | | | | | | |
| Madhya Pradesh (84.3) | Mar-May, 3 months (2021 vs 2018-19) | 268 | 176,009 | 52.3% | 89% | 51% |
| Haryana (29.3) | April-May, 2 months (2021 vs 2018-19) | 74 | 34,816 | 113.7% | 100% | 81% |
| Andhra Pradesh (52.5) | Apr-Jun, 3 months (2021 vs 2018-19) | 216 | 102,650 | 110.9% | 100% | 80% |
| Tamil Nadu (82.3) | March-May, 3 months (2021 vs 2018-19) | 203 | 152,965 | 32.9% | 100% | 91% |
| Gujarat (69.5), DC | Mar-May, 2.3 months (2021 vs 2020) | 124 | 85,775 | 44.4% | 100% | 101% |
| Odisha (46.5) | Jan-Jun, 5.6 months (2021 vs 2015-19) | 191 | 163,535 | 16.9% | 100% | 95% |
| Kerala, (35.3) | Apr-May, 2 months (2021 vs 2019) | 48 | 46,551 | 3.5% | 100% | 84% |
| | ***Sub-totals and medians*** | **1,125** | **762,300** | **44.4%** | **100.0%** | **84.3%** |
| **Cities (Wave 2)** | | | | | | |
| Hyderabad, TL (9.7) | Apr-May, 2 months (2021 vs 2016-19) | 20 | 8,614 | 135.9% | 97% | 102% |
| Bengaluru, KN (8.5) | Apr-May, 2 months (2021 vs 2019) | 22 | 8,642 | 160.3% | 100% | 124% |
| Chennai, TN (8.7) | Mar-Apr, 2 months (2021 vs 2018-19) | 18 | 9,723 | 84.2% | 100% | 100% |
| Mumbai, MH (18.4) | Mar-May, 3 months (2020 vs 2017-19) | 30 | 24,240 | 25.0% | 100% | 88% |
| Kolkata, WB (14.1) | Mar-Apr, 2 months (2021 vs 2018-19) | 15 | 14,996 | 0.0% | 100% | 69% |
| | ***Sub-totals and medians*** | **106** | **66,214** | **84.2%** | **100.0%** | **99.8%** |

**Table S5 - Civil registration uncounted deaths in major Indian states**

| State/union territory | SRS-UN deaths (0000) | Civil registration death 2017-19 (000) | | | Medically certified deaths 2017-18 (000) | % Registered but not medically certified | | % uncounted deaths at | | |
|---|---|---|---|---|---|---|---|---|---|---|
| | | Rural urban together | Urban | Female | | All India | Urban | Both sexes at civil registration | Females at Civil registration# | At the medical certification |
| India | 10,008.6 | 7,018 | 2,983 | 2,846 | 1,355 | 80.7 | 54.6 | 29.9 | 59.3 | 86.5 |
| Andhra Pradesh/ Telangana | 711.5 | 558.7 | 231.2 | 223.7 | 106.2 | 81.0 | 54.1 | 21.5 | 80.5 | 85.1 |
| Assam | 207.2 | 148.9 | 41.3 | 59.7 | 32.1 | 78.5 | 22.3 | 28.2 | 64.5 | 84.5 |
| Bihar | 756.2 | 278.3 | 72.3 | 105.9 | 19.3 | 93.1 | 73.3 | 63.2 | 57.6 | 97.4 |
| Chhattisgarh | 250.5 | 180.3 | 65.6 | 75.4 | 33.9 | 81.2 | 48.3 | 28.0 | 51.9 | 86.5 |
| Delhi | 83.1 | 142.3 | 125.6 | 54.6 | 84.8 | 40.4 | 32.5 | | 34.1 | |
| Gujarat | 441.0 | 428.0 | 197.7 | 172.5 | 84.4 | 80.3 | 57.3 | 2.9 | | 80.9 |
| Haryana | 208.9 | 183.2 | 82.7 | 67.6 | 30.4 | 83.4 | 63.2 | 12.3 | 85.9 | 85.4 |
| Jharkhand | 220.6 | 112.8 | 45.1 | 45.9 | 5.0 | 95.5 | 88.8 | 48.9 | 60.4 | 97.7 |
| Karnataka | 639.0 | 491.3 | 218.8 | 192.4 | 147.9 | 69.9 | 32.4 | 23.1 | 80.5 | 76.9 |
| Kerala | 279.3 | 264.1 | 98.8 | 118.4 | 29.2 | 88.9 | 70.4 | 5.4 | 73.3 | 89.5 |
| Madhya Pradesh | 704.0 | 429.4 | 168.8 | 164.0 | 37.4 | 91.3 | 77.8 | 39.0 | 53.0 | 94.7 |
| Maharashtra | 873.4 | 669.6 | 393.0 | 282.8 | 207.3 | 69.0 | 47.3 | 23.3 | 68.9 | 76.3 |
| Odisha | 349.1 | 331.5 | 86.8 | 141.9 | 39.7 | 88.0 | 54.3 | 5.0 | 65.6 | 88.6 |
| Punjab | 196.9 | 212.9 | 100.9 | 87.8 | 34.0 | 84.0 | 66.3 | | 6.2 | 82.7 |
| Rajasthan | 582.0 | 439.8 | 145.7 | 156.2 | 56.7 | 87.1 | 61.1 | 24.4 | 55.5 | 90.3 |
| Tamil Nadu | 611.9 | 596.1 | 300.6 | 241.7 | 248.7 | 58.3 | 17.3 | 2.6 | | 59.3 |
| Uttarakhand | 107.4 | 49.4 | 21.1 | 18.9 | 4.1 | 91.6 | 80.4 | 54.0 | 57.5 | 96.2 |
| Uttar Pradesh | 1,865.1 | 807.5 | 282.6 | 352.0 | 43.4 | 94.6 | 84.6 | 56.7 | 47.6 | 97.7 |
| West Bengal | 675.9 | 495.1 | 218.5 | 205.0 | 47.7 | 90.4 | 78.2 | 26.8 | 65.8 | 92.9 |
| Jammu & Kashmir | 66.9 | 40.3 | 14.4 | 17.6 | NA | NA | NA | 39.8 | 31.0 | NA |

Notes:

1. ** - SRS-UN death estimates: SRS death rates applied to United Nations estimated population and then adjusted to UN estimated deaths. SRS death rates are under reported. In most civil registration completed states, registered deaths are closer to these estimated deaths.
2. # - numerator for % un-accounted female deaths at civil registration was the difference of SRS-UN estimated female deaths and the female civil registration deaths. SRS-UN estimated female death not shown in this table.

Sources of data: SRS vital Statistics reports, Medical certification of Causes of Death (MCCD), Vital statistics of India civil registration system, World Population Prospectus (WPP) of United Nations Population Division.

**Figure S1 - Reporting PCR Positive rates in population and undocumented deaths at civil registration in Indian states**

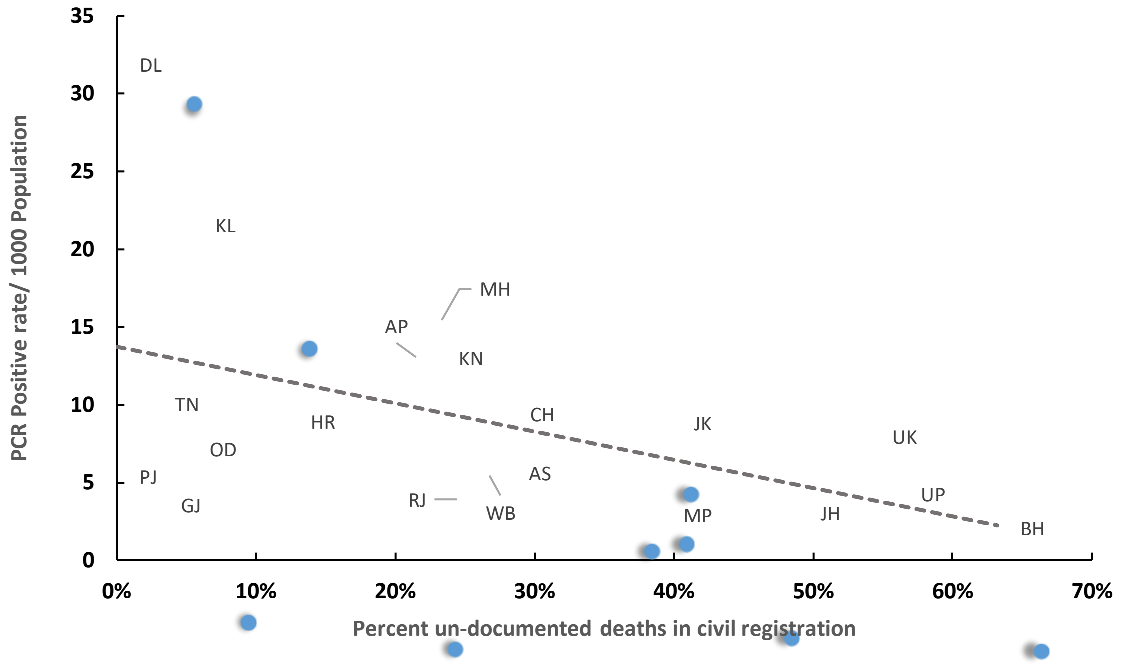


Notes: PCR positive rates for Apr-Dec 2020. Percent un-documented deaths were calculated using the difference between average civil registration deaths for 2017-19 and the expected deaths calculated for each state from SRS death rates adjusted for UN death estimates. The Pearson correlation between PCR positivity rate and the percent un-documented deaths in civil registration was -0.5.

Abbreviations: AP - Andhra Pradesh/Telangana, AS - Assam, BH - Bihar, CH - Chhattisgarh, DL - Delhi, GJ - Gujarat, HR - Haryana, JH - Jharkhand, JK - Jammu & Kashmir, KL - Kerala, KN - Karnataka, MH - Maharashtra, MP - Madhya Pradesh, OD - Odisha, PJ - Punjab, RJ - Rajasthan, TN - Tamil Nadu, UK - Uttarakhand, UP - Uttar Pradesh, WB - West Bengal.

**Figure S2 – Un-counted at civil registration and non-medically certified deaths in India 2019**. Out of the 10 million annual deaths in India, 3 million deaths are un-accounted at civil registration and 8.5 million are not medically certified

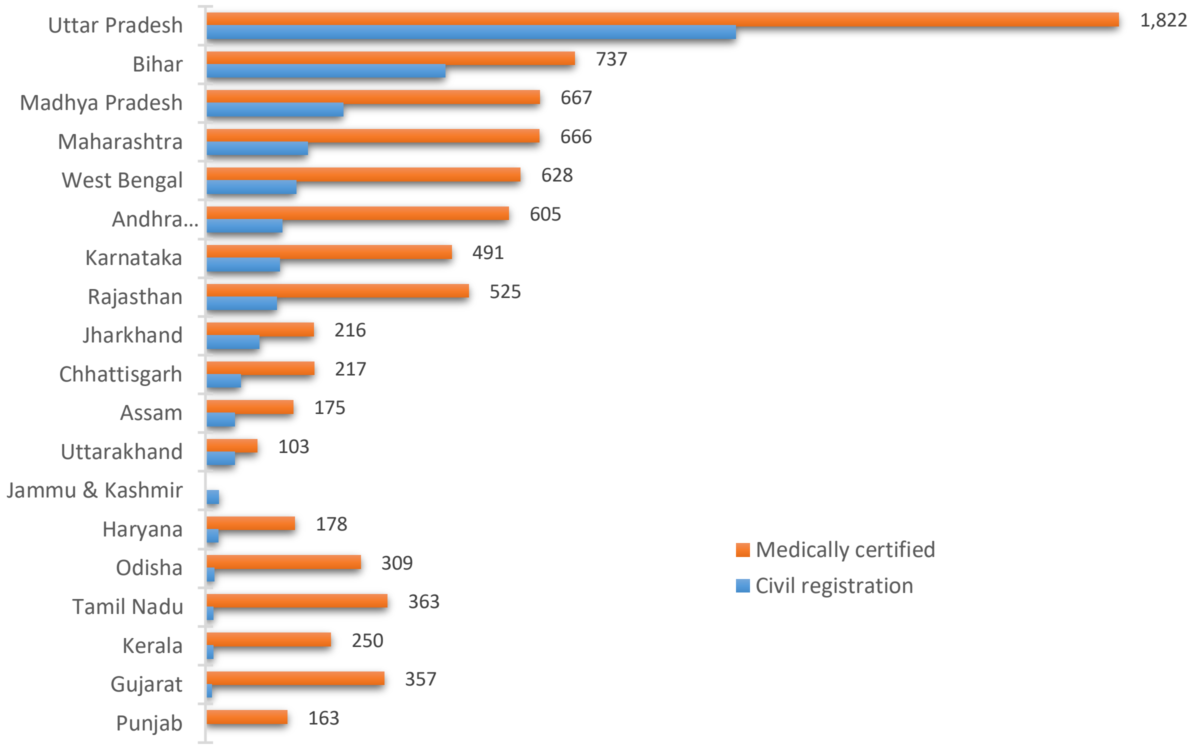
